# Projecting the Spread of COVID19 for Germany

**DOI:** 10.1101/2020.03.26.20044214

**Authors:** Jean Roch Donsimoni, René Glawion, Bodo Plachter, Klaus Wälde

## Abstract

We model the evolution of the number of individuals that are reported to be sick with COVID-19 in Germany. Our theoretical framework builds on a continuous time Markov chain with four states: healthy without infection, sick, healthy after recovery or after infection but without symptoms and dead. Our quantitative solution matches the number of sick individuals up to the most recent observation and ends with a share of sick individuals following from infection rates and sickness probabilities. We employ this framework to study inter alia the expected peak of the number of sick individuals in a scenario without public regulation of social contacts. We also study the effects of public regulations. For all scenarios we report the expected end of the CoV-2 epidemic.

We have four general findings: First, current epidemiological thinking implies that the long-run effects of the epidemic only depend on the aggregate long-run infection rate and on the individual risk to turn sick after an infection. Any measures by individuals and the public therefore only influence the dynamics of spread of CoV-2. Second, predictions about the duration and level of the epidemic must strongly distinguish between the officially reported numbers (Robert Koch Institut, RKI) and actual numbers of sick individuals. Third, given the current (scarce) medical knowledge about long-run infection rate and individual risks to turn sick, any prediction on the length (duration in months) and strength (e.g. maximum numbers of sick individuals on a given day) is subject to a lot of uncertainty. Our predictions therefore offer robustness analyses that provide ranges on how long the epidemic will last and how strong it will be. Fourth, public interventions that are already in place and that are being discussed can lead to more and less severe outcomes of the epidemic. If an intervention takes place too early, the epidemic can actually be stronger than with an intervention that starts later. Interventions should therefore be contingent on current infection rates in regions or countries.

Concerning predictions about COVID-19 in Germany, we find that the long-run number of sick individuals (that are reported to the RKI), once the epidemic is over, will lie between 500 thousand and 5 million individuals. While this seems to be an absurd large range for a precise projection, this reflects the uncertainty about the long-run infection rate in Germany. If we assume that Germany will follow the good scenario of Hubei (and we are even a bit more conservative given discussions about data quality), we will end up with 500 thousand sick individuals over the entire epidemic. If by contrast we believe (as many argue) that once the epidemic is over 70% of the population will have been infected (and thereby immune), we will end up at 5 million cases.

Defining the end of the epidemic by less than 100 newly reported sick individuals per day, we find a large variation depending on the effectiveness of governmental pleas and regulations to reduce social contacts. An epidemic that is not influenced by public health measures would end mid June 2020. With public health measures lasting for few weeks, the end is delayed by around one month or two. The advantage of the delay, however, is to reduce the peak number of individuals that are simultaneously sick. When we believe in long-run infection rates of 70%, this number is equally high for all scenarios we went through and well above 1 million. When we can hope for the Hubei-scenario, the maximum number of sick individuals will be around 200 thousand “only”.

Whatever value of the range of long-run infection rates we want to assume, the epidemic will last at least until June, with extensive and potentially future public health measures, it will last until July. In the worst case, it will last until end of August.

We emphasize that all projections are subject to uncertainty and permanent monitoring of observed incidences are taken into account to update the projection. The most recent projections are available at https://www.macro.economics.unimainz.de/corona-blog/.

## 1 Introduction

There is no need to stress the importance of the Coronavirus disease (COVID-19) for public health, economic consequences and well-being of individuals. Yet, there is a need for more knowledge about objective information and reliable predictions of how the pandemic will likely evolve in the months to come. How large is the risk of any individual to get infected on a particular day or within a week? How does this risk change over time? How large is the expected number of individuals ever infected and how large is the number of individuals infected at any point in time? The latter question is the main concern of the public, politicians and health practitioners to ensure sufficient provision of health services. Finally, what are the effects of policy interventions that are already implemented and that are being discussed?

This paper starts by describing what we know quantitatively about the spread of the Corona epidemic in the Chinese province of Hubei and in South Korea. This offers insights from two episodes of epidemics that seem to be coming to an end and that help in making predictions for other countries. We then compare the time-series evidence from Hubei and South Korea with cross-sectional information in European countries.

Section 3 develops an epidemiological Markov model. We start from individuals that can be in four states: healthy (the initial state, state 1), infected and sick (state 2), infected and recovered or infected and never having displayed symptoms (state 3) and dead (state 4). Individuals move between these states with endogenous transition rates that depend on population characteristics. This model allows to make predictions, inter alia, about all the quantities of interest that appeared in the questions raised initially.

We apply this model to Germany. We choose parameter values such that the time series of the number of sick (and reported) individuals in Germany since 24 February is matched by our model. Our data source is the Robert Koch Institute (RKI, 2020). In order to predict the future evolution, we build on various evidence from epidemiology like individual sickness probability and long-run infection rates of the population as a whole. We emphasize the scarce medical and statistical knowledge on these parameters. Any predictions therefore must be subject to large variations. We therefore undertake robustness checks with respect to our central parameters.

Given our calibration for the epidemic without public intervention, we predict the number of sick individuals at each point in time. We compute when the peak will be reached and whether medical services are in sufficient supply. We also undertake counterfactual analysis to understand when drastic policy measures (like shutting down educational institutions or canceling big public events) are most effective.

There is an exploding literature on these issues, especially in medical science. The classic large-scale spatial model is described in Balcan et al. (2010). It is applied in Chinazzi et al. (2020) to study the effect of a travel ban on China and the world. Akbarpour and Jackson (2018) study a general model of infection, but cannot provide applications to the current epidemic. The widely discussed study by Ferguson et al. (2020) focuses on the US and the UK.^2^ An insightful study on the usefulness of isolation strategies for COVID-19 based on a stochastic transition model similar to ours is by Hellewell et al. (2020). Wilder-Smith et al. (2020) discuss the similarities and differences between the SARS 2003 (severe acute respiratory syndrome) epidemic and COVID-19. One big issue is the large number of “quiet infections” in COVID-19 as opposed to SARS 2003, an aspect we will capture in our model below.

The analyses which come closest to ours are two notes by the Deutsche Gesellschaft für Epidemiologie (DGfE, 2020) and by an der Heiden and Buchholz (2020). DGfE (2020) offer predictions based on a model similar to ours (so called SEIR models, see e.g. Anderson et al., 1992). We present our model in detail, including the stochastic foundation, and discuss the implications of the modelling assumptions.^3^ Modelling assumptions turn out to be crucial for evaluating public policy measures. We also implement in detail the effects of policy measures and discuss the trade-offs with respect to length and strength of the evolution of the epidemic. The match of the model with the data also differs. While DGfE (2020) seem to use summary statistics for their calibration, we determine the parameters of our model by fitting our model predictions to the data. This offers the advantage that we can update our predictions whenever new data is available.

The study by an der Heiden and Buchholz (2020) is, to the best of our knowledge, the most elaborate study at this point for Germany. We share their belief that mathematical models help in studying the effects of policy measures. Many parameters of their analysis are derived from knowledge on the epidemic in China and especially the city Shenzen. We calibrate many of our central parameters such that the model fits the observations in Germany. We also share with them the approach of using SEIR models. Our paper emphasises the implications of the fundamental model assumptions for the prediction. We find below that the evaluation of policy measures crucially depends on these model assumptions. We make these model assumptions explicit in our paper and discuss the effects of variations in these assumptions.

Why should economists work on an epidemic? The economic costs of COVID-19 are huge and seem to be larger than those of the financial crisis starting in 2007.^4^ This paper provides a model that allows to understand the spread of the disease. We employ a model in the tradition of search and matching models originating from Diamond (1982), Mortensen (1982) and Pissarides (1985). The basic structure of these models (continuous time Markov chains) is identical to epidemiological models (as becomes clear very quickly when reading overviews like e.g. Hethcote, 2000). Compared to standard search and matching models, we do allow for four states, however. We extend the typical matching framework from a technical perspective by taking the stochastic nature of transitions and their prediction for the probability to be in various states into account. These probabilities are described by forward Kolmogorov equations. As we work with a discrete number of states, we obtain an ordinary differential equation system.^5^ The conclusion discusses various next steps that should be undertaken to understand economic consequences of COVID-19 better. We believe that our framework can form the basis for an understanding of economic costs of the pandemic.

### 2 Data

This section describes what we know quantitatively about the spread of reported infections. We start with Hubei and its capital Wuhan in China. We afterwards look at South Korea and very briefly at Japan. We focus on the evolution of the number of individuals that were ever reported to be sick during the current epidemic. This number, by construction, can only rise. We chose Hubei and South Korea as the epidemics seem to come to an end in these countries. Given the huge uncertainty about long-run infection rates and individual risk to get sick after an infection with CoV-2, we consider the long-run ratios of sick reported individuals to the overall population a benchmark that allows us to judge the credibility of our predictions for Germany. We are aware that we compare very different countries with very different cultural and political habits and medical systems. For reference, we also look at a cross section of infections for European countries. See the appendix for a description of data sources.^6^

#### Hubei

Since the onset of the outbreak in the Hubei province in China, the number of confirmed cases has risen dramatically, peaking at 67,801 (WHO Situation Reports, 2020).^7^ Reporting began in January 2020 and we note that in mid-February, the WHO changed its classification methodology by reporting both clinically and lab confirmed cases rather than only lab confirmed cases. In terms of cumulative numbers, the total reported cases of infected individuals rose continuously since January, since there have been continuous influxes of new cases and very little communication about individuals transitioning back to being healthy.

When reporting began, the ascension of new cases seemed to follow an exponential curve, with the number of new cases constantly increasing, however in early February, this trend stopped. Indeed, another trend then took over following a more S-shaped pattern, i.e. that of a sigmoidal function. The total number of reported cases over time is shown in Figure 1. The figure shows a clear explosive trend at the start of the outbreak before slowing down and approaching what appears to be an asymptote around 70,000.

**Figure 1.**
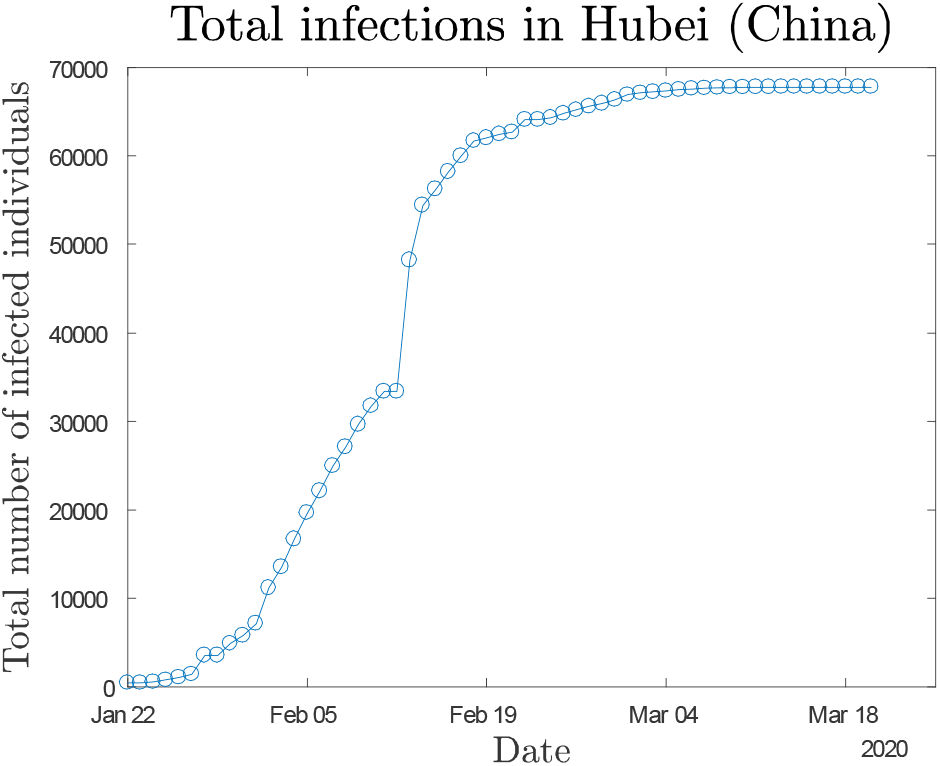
Total cases in Hubei seem to converge to a constant

To make this comparable to other countries, we take the population of around 59.17 million individuals in Hubei into account. This implies an infection rate of about 0.11% or 1 person in 873 being infected in the long-run. While this is only one province, it does provide us with a reference point for what to expect in other regions of the world.

#### South Korea and Japan

Soon after China, South Korea, Japan, and many other countries all started seeing a rise in their number of confirmed cases. However, these countries have had very different experiences when it comes to infection outbreaks. Indeed, while South Korea has seen the number of confirmed cases rise to 9,241, Japan only has 1,307 confirmed cases (or 14% that of South Korea). The question remains to know whether these countries are also experiencing a convergence to a stable number of (cumulative) confirmed cases. Figure 2 shows the cumulative number of confirmed cases in South Korea (left) and Japan (right).

**Figure 2.**
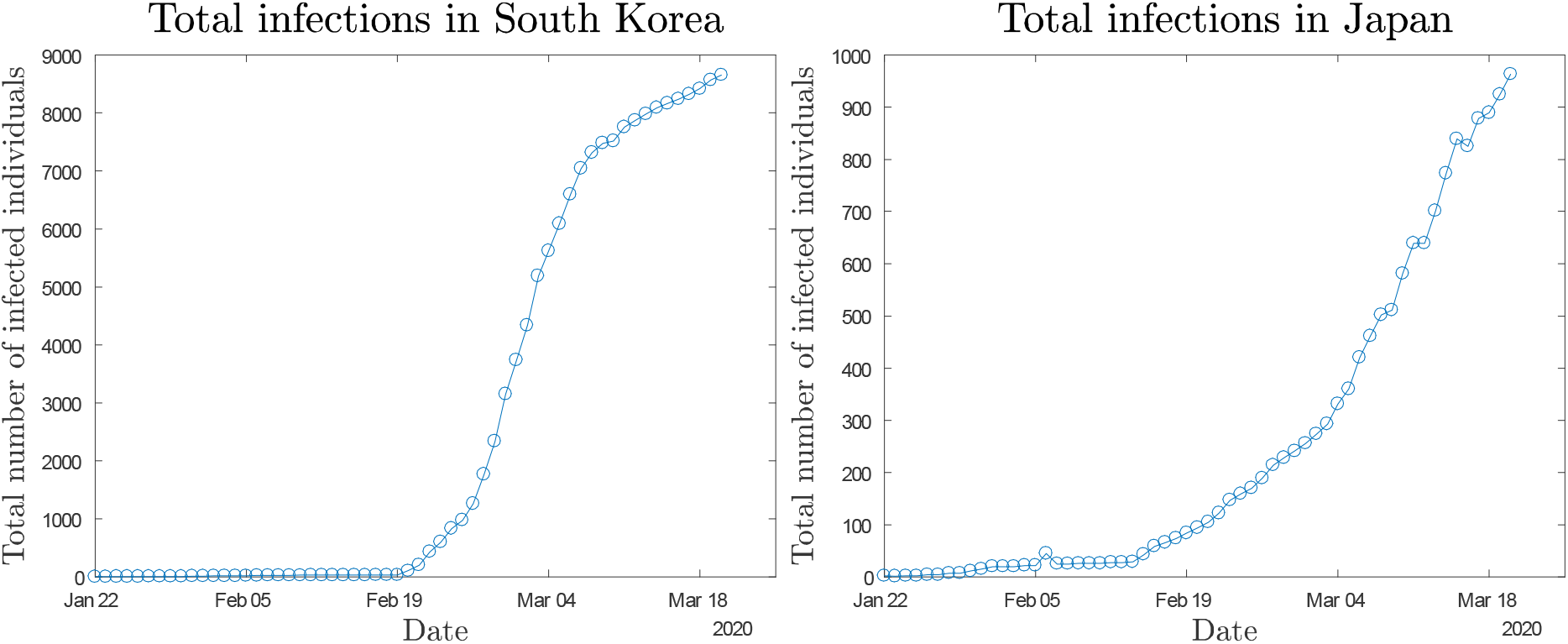
Total cases in South Korea exhibit a similar pattern to the Hubei province

The data from South Korea shows a reduction in the slope of the curve for total confirmed cases, indicating a significant drop in the number of new infections reported. Assuming the tendency continues, this would point toward South Korea progressively converging to a constant, similarly to the Hubei province in China. Japan on the other hand (as well as all other countries at this point) does not appear to slow down significantly. Relating South Korean infections to population size, we get a long-run infection rate of 1 in 5,337, with a peak number of confirmed cases of ca. 9,606.

#### Europe

In Europe, however, the situation continues to deteriorate, with new cases being reported by authorities on a daily basis numbering in the thousands (WHO Situation Reports, 2020). However, bearing in mind the lessons learned from the case of the Hubei province, and to a lesser extent from the case of South Korea, this current period of explosive growth should, in theory, be followed by a slow tapering leading to a plateauing of the total number of cases. We report in Table 1 the current infection rates in some European countries. The sample below represents 58% of the total EU population, including Norway and Switzerland. This sample is by no means intended to be representative and is for illustrative purposes only by considering some of the most affected economies.

**Table 1.**
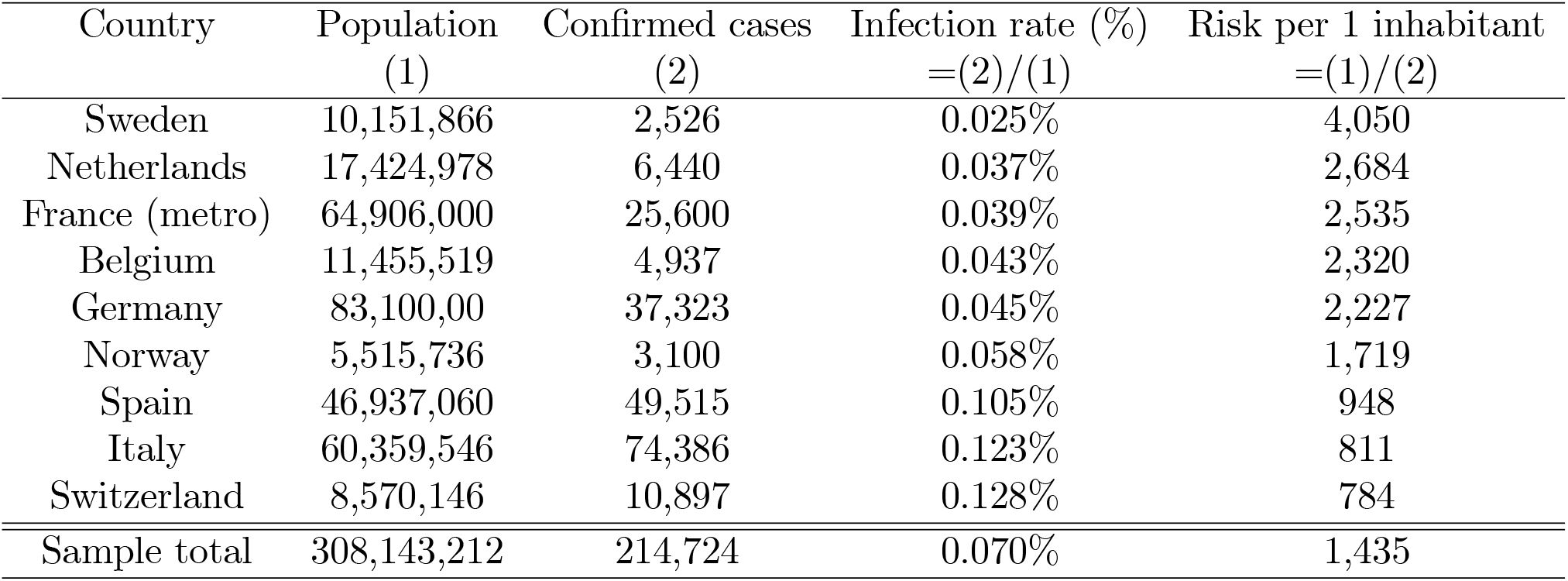
Current infection rates and risks in a sample of European countries

As we can see from Table 1, the degree of infection per country varies from 1 in 4,050 to 1 in 784, a fivefold ratio. Other countries fare better, the UK for example has a risk of 1 in 6,914, however we can clearly see that these figures are not yet as low as those in Hubei, with the exception of Italy and Switzerland, which are now below the 1 in 873 rate observed in the Chinese province, and Spain, which is approaching it fast. If this case is to be repeated, then things will likely worsen before they improve. However, with measures taken by governments, and progress being made on the medical front for an effective vaccine, there is still room for governments to curb these rates despite the rapid rise in cases.

## 3. The model

### 3.1 The structure

The model builds on a continuous time Markov chain with 4 states. We look at health of an individual *i* that can be in four states *S* = {1, 2, 3, 4}. Healthy without infection (*s* = 1), sick (*s* = 2), dead (*s* = 3), healthy with or after infection (*s* = 4). State 1 is the initial state for all individuals, thus *s*_*i*_ (0) = 1 for all *i* = 1…*N* where *N* is initial population size.^8^ There are individual transition rates λ_*rs*_ (with *r* ∈ *S* and *s* ∈ *S*) between these states as illustrated in figure 3.

**Figure 3.**
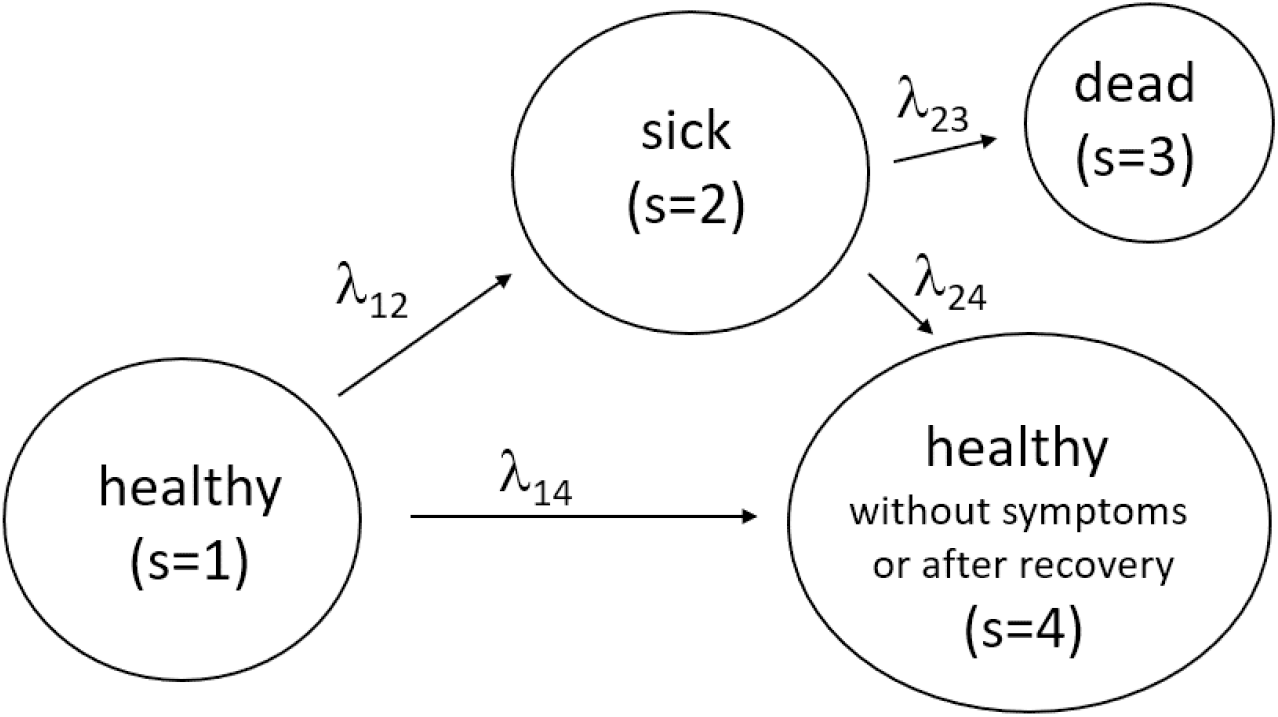
Transitions between the state of health (initial state), sickness, death and health despite infection or after recovery

### 3.2 Transition rates

Starting with the transition from being healthy to sick, each individual in state 1 has a certain average number of social contacts per day. We capture this by an exogenous arrival or contact rate *a*. Once a contact takes place, there is a certain probability that the contact is with an individual that can infect the healthy individual. If we assumed random contacts with healthy and sick individuals, this probability would be (*N*_2_ (*t*)+ *ηN*_4_ (*t*)) */* (*N*_1_ (*t*)+ *N*_2_ (*t*)+ *N*_4_ (*t*)), where *N*_*s*_ (*t*) is the (expected) number of individuals in state *s* at *t*. This probability assumes that all *N*_2_ (*t*) individuals that are sick can infect healthy individuals and a constant and exogenous share *η* of healthy individuals in state 4 can also infect healthy individuals from state 1. This parameter *η* captures the idea of “quiet infections” as emphasized e.g. by Wilder-Smith et al. (2020). As opposed to SARS 2003 (or the common flu), individuals infected with the CoV-2 do not necessarily display symptoms but can nevertheless infect others. There is also first evidence (Xing et al., 2020) that even recovered individuals (i.e. those that came from state 2) can carry the CoV-2, also supporting our use of a positive *η*.

In practice, individuals do not encounter sick individuals with this probability being equal to the share of sick individuals in society as sick individuals might stay at home or behave differently (not shaking hands or similar) than non-sick individuals. We do assume, however, that the probability π (*t*) to encounter a sick individual rises in *N*_2_ (*t*) and *ηN*_4_ (*t*) and falls in *N*_1_ (*t*), i.e. π (*t*) = π (*N*_2_ (*t*), *ηN*_4_ (*t*), *N*_1_ (*t*)) with ∂π*/*∂*N*_2_ *>* 0, ∂π*/*∂ (*ηN*_4_) *>* 0 and ∂π*/*∂*N*_1_ *<* 0.9 These contacts can lead to infections. Hence, we have an individual sickness rate of λ_12_ (*t*) ≡ *a*π (*t*). Finally, when there are *N*_1_ (*t*) healthy individuals, the aggregate flow from state 1 to state 2 is φ_12_ (*t*) ≡ *a*π (*t*) *N*_1_ (*t*).^10^

A further property an infection rate should reflect is the fact emphasized by many virologists and epidemiologists that once an epidemic is over, around two thirds of the entire population will be infected. This includes individuals that had symptoms at some point and asymptomatic cases. We capture this fact by introducing the infection rate which is simply the ratio of the number of infected individuals (sick and in state 2 or without symptoms in state 4) to individuals that are alive,

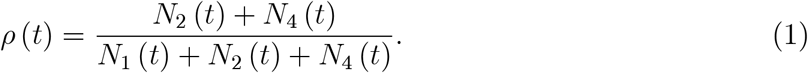

The infection rate is zero initially at *t <* 0. Illustrated for Germany in a somewhat too simple way but perfectly fine for our projection, ρ = 0 on 23 February 2020 (and before) when the number of reported sick individuals was zero. On 24 February 2020, a number of *N*_2_ (0) = 16 are introduced into the system and infections and sickness start occurring. The sickness rate λ_12_ (*t*) is zero also in the long-run when this infection rate equals the long-run infection rate, which we denote by ρ. This captures the above mentioned fact that some individuals will never be infected during the epidemic and will always remain in state 1, lim _*t*→∞_*N*_1_ (*t*) *>* 0. Summarizing our discussion, an example for the individual sickness rate that captures insights from epidemiology and virology would be

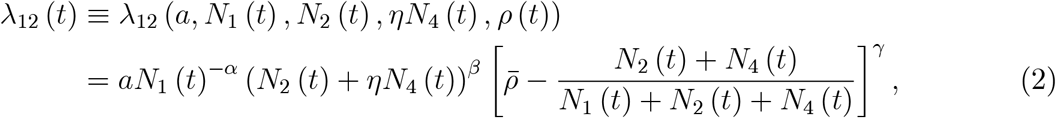

where 0 *<* α, β, γ *<* 1 allows for some non-linearity in the process and *a >* 0. The first term *N*_1_ (*t*)^−α^ captures the idea that more healthy individuals reduce the individual sickness rate. The second term (*N*_2_ (*t*)+ *ηN*_4_ (*t*))^β^ increases the sickness rate when there are more infectious individuals. The third term in squared brackets makes sure that the arrival rate is zero when a share ρ of society is sick (state 2) or healthy after infection (state 4). The sickness rate satisfies “no sickness without infected individuals”, 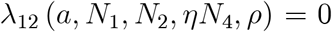 and “end of spread at sufficiently high level”, 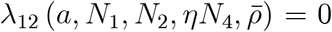 In between these start- and endpoints, the infection rate will first rise and then fall. This specification makes sure that in the long run a share of around 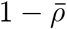 will not have left state 1, i.e. will never have been infected.^11^

As is widely documented and reported (see e.g. Nishiura et al., 2020), there is also a flow of individuals that are infected but do not display any symptoms. We denote this flow by φ_14_ (*a, N*_1_ (*t*), *N*_2_ (*t*), *ηN*_4_ (*t*)) and the corresponding individual rate by λ_14_. The arguments can be rationalized in the same way as the flows into sickness.

Is it possible to obtain data for λ_14_? One would need a cohort of inhabitants of a region (and not just those that contact medical services to get tested) to understand the share of individuals that are infected but do not display symptoms. Good data is not available (let alone time series) and might never be available, given the currently still large costs per individual tests (between 80 Euro and 120 Euro). We therefore assume that a constant share *r* of infected individuals display symptoms. This is a quantity many epidemiologists also work with. Hence, the transition rate λ_14_ is related to λ_12_ in the following sense,

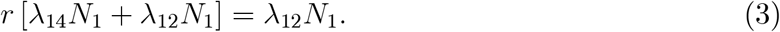

In words, the outflow of newly infected, λ_14_*N*_1_ + λ_12_*N*_1_ in squared brackets on the left-hand side, times the share *r* of individuals that show symptoms after infection gives the flow into sickness, λ_12_*N*_1_. We can therefore compute the transition rate from the state of being healthy (*s* = 1) to the state of healthy again or no symptoms (*s* = 4) as

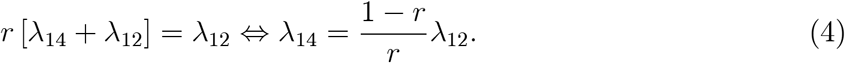

Finally, there is an individual mortality rate and a recovery rate. Both rates depend on many individual characteristics. At the aggregate level, they rise in the stock of sick individuals. We specify them as φ_23_ (*N*_2_ (*t*)) and φ_24_ (*N*_2_ (*t*)) with corresponding rates λ_23_ and λ_24_, respectively.^12^ For our analysis below, we will assume that both rates are constants. Concerning recovery, we are aware that it strongly varies across age (e.g. Guan et al., 2020) and that recovery is not a state that is identical for each individual. As we abstract from ex-ante heterogeneity, we assume a recovery rate that is identical across individuals and that it takes on average *n*_rec_ days until recovery. Given the exponential distribution of duration in each state, this means that 1*/*λ_24_ = *n*_rec_ or

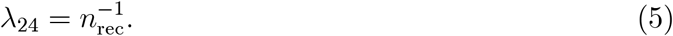

Death is an absorbing state. We also consider the final healthy state to be absorbing, i.e. we abstract from relapses.

### 3.3 Probabilities and means

#### The evolution of probabilities, shares and expected numbers

We can express the transition across health states of an individual by a stochastic differential equation similar to the transition across states of employment (see e.g. appendix *B*.1 to Bayer et al., 2019 or Khieu and Wälde, 2020). We can also directly write down ordinary differential equations for an individual to be in a certain state *s* at *t*. Let us denote this probability by *p*_*s*_ (*t*) =Prob(*s*_*i*_ (*t*) = *s*|*s*_*i*_ (0) = 1), i.e. as the probability to be in state *s* at *t* conditional on having started in state 1 at *t* = 0. The ODEs have the following general form (see Ross, 1993 for background),

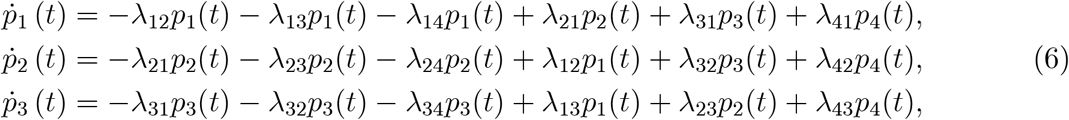

and are completed by the identity *p*_4_ (*t*) = 1 − *p*_1_ (*t*) − *p*_2_ (*t*) − *p*_3_ (*t*). For brevity, individual transition rates are represented by λ_*rs*_. We stress that these equations hold for arrival rates that can take any functional form with arguments *N*_1_, *N*_2_, *N*_3_ and *N*_4_.

As death is an absorbing state, λ_3*i*_ = 0 for all *i*. Given the assumption that infected means being immune to further infection and that the mortality rate is negligible in symptom-free states, we also set λ_21_ = λ_41_ = λ_13_ = λ_43_ = 0. Our final structure for the evolution of individual probabilities therefore reads

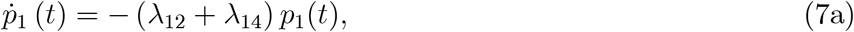

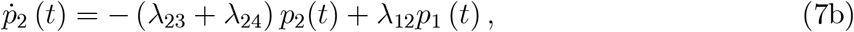

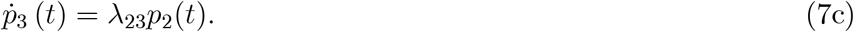

A standard law of large numbers tells us that individual probabilities also give shares in the population. Hence, when we solve this system, we can think of the time paths of *p*_*s*_ (*t*) also as the shares of society in the different states. When we compute the expected number of individuals in state *s* at *t*, denoted by *N*_*s*_ (*t*), we would also obtain an identical ODE system where the probabilities are replaced by the expected numbers.^13^ As the interest of our analysis lies in the (expected) number of individuals in certain states (are there sufficiently many beds in hospital for severe cases?), we will also refer to the variable *p*_*s*_ (*t*) as the (expected) number *N*_*s*_ (*t*) of individuals.

#### The probability to get sick per day

For our quantitative predictions it will be useful to present the probability for an individual to get sick over a period of one day or one week. So far, we only talked about rates λ_*rs*_. We denote the probability to get sick over a period of one day, starting in *t*, by 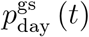 Thisprobability is one minus the probability to stay healthy. The probability to stay healthy is, given the exponential distribution of duration in a state, a simple exponential function of the transition rate. We therefore obtain

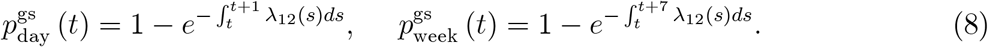

The second probability 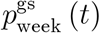 is the probability to get sick between today in *t* and the next 7 days to come. We will plot them below to provide a measure of how risky social interactions are during the evolution of the epidemic.

### 3.4 The number of individuals who get and ever got sick and deaths

As recovery is a process that enters additional measurement problems, a more reliable data source for our calibration purposes is the number of individuals that have been reported to be sick since the onset of the epidemic. This is also the time series that is typically reported by countries and regions (such as those reported by the RKI in Germany or the WHO more generally). In our model, the corresponding number follows from the inflows from state 1 to 2 and is given by

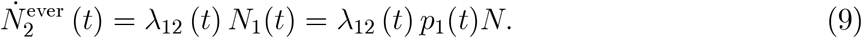

When we are interested in the number of individuals that are newly reported to be sick at some point in time *t* (incidences at *t* in short), we should not employ *N*_2_ (*t*). The change of the latter, described in (7b), is determined by the newly reported (inflow) minus the outflow, i.e. those that recovered (and those who died). Hence, when data provides incidences on a given day *t*, we need to compare this with the inflow between yesterday *t* 1 and *t*. Hence, our theoretical counterpart to incidences is

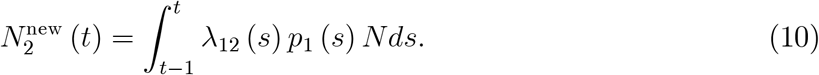

Finally, the number *N*_3_ (*t*) of deaths from Corona by *t* is determined by the constant death rate λ_23_ applied to those that are currently sick,

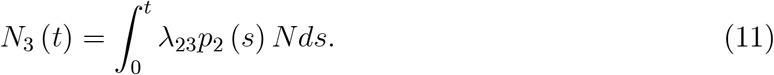

### 3.5 Model properties

Before we calibrate our model, we briefly discuss its theoretical predictions. This illustrates the plausibility of our assumptions and also the flexibility of the model. Our model consists of the ODE system (7) where we replace the probabilities *p*_*s*_ (*t*) by the expected number *N*_*s*_ (*t*) as described in footnote 13. The sickness rate λ_12_ is from (2), the other arrival rates λ_14_ and λ_24_ are from (4) and (5), respectively. The death rate λ_23_ is a constant. We start the solution of our model with 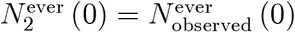 We set *N*_3_ (0) = *N*_4_ (0) = 0 and 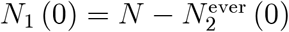.

One can easily understand the plausible structure of our ODE system (7). The number *N*_1_ (*t*) of healthy individuals in state 1 (i.e. the probability or share *p*_1_ (*t*)) can only fall. Individuals either get sick with rate λ_12_ or get infected without symptoms with rate λ_14_. Both transitions imply an outflow from state 1. The same is true, mutatis mutandis, for state 3 and 4: For state 3, there are only inflows from state 2 implying that *N*_3_ (*t*) increases over time. State 4 only experiences inflows from state 1 and 2 implying that *N*_4_ (*t*) increases over time. The number *N*_2_ (*t*) of sick individuals can rise or fall, depending on the difference between inflows with λ_12_ and outflows with λ_23_ or λ_24_.

In the long-run, i.e. when the epidemic is over, individuals will be distributed across states 1, 3 and 4. Some individuals will never get infected. They therefore remain in state 1 all throughout the epidemic. This finding follows from the fact that the sickness rate λ_12_ from (2) falls to zero when the infection rate ρ (*t*) from (1) equals its long-run value 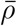 Note that λ_12_ = 0 implies from (4) that λ_14_ = 0 as well, hence outflows from 1 to 4 end as well. The ratio 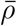 is a widely accepted quantity in epidemiology and will be quantified below. Some individuals that turn sick and are in state 2 die. They end up in state 3 in the long-run. Most individuals will be in state 4 in the long-run, either after having transitioned through state 2 or directly from state 1.

The model also displays a long-run property that is extremely useful to judge its quantitative property in a very simple way. With a population of *N* individuals and a long-run infection rate 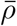 discussed after (1), the number of individuals that will have been infected is 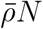 When a share *r* of all infected turns sick, the long-run number of individuals that were sick at some point, described in (9), is

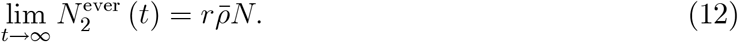

The long-run number of reported sick individuals therefore only depends on the long-run infection rate 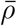 and the share *r* of individuals that turn sick *and* are reported. We will exploit this property when we discuss the effects of policy measures below.

## 4 Calibration

### 4.1 Data

Our most important data source is the number of individuals that is reported to be sick. We obtain these outflows from state 1 to 2 from the Robert Koch Institute (RKI, 2020). These data fix the left-hand side of our time path for the number 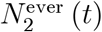 from (9) of individuals that are ever reported to be sick. To fix notation, *t* = 0 stands for the 24th February 2020 and *T* is the day of our final observation.

It seems reasonable to assume that parameters are not constant throughout the entire epidemic. Most centrally, the contact rate *a* employed in the sickness rate (2) will change as a function of governmental regulations. The first major effects consisted in cancelling sports events (14 March) and closing schools (16 March). Other measures followed.^14^ Given a median incubation of 5.2 days (Linton et al., 2020, Lauer et al., 2020) and a certain delay between feeling symptoms, contacting a doctor and getting reported at RKI (say, 2-3 days), we employ *T* = 21 March 2020 as our last observation to fix the contact rate *a*.

Data to compute the death rate from Corona are also taken from the RKI (2020). We finally employ various sources, to be discussed in what follows, to fix the share *r* of infected individuals that turn sick and the long-run infection rate 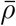.

### 4.2 Targets and parameters

#### Parameters

Some model parameters are straightforward to be fixed. Concerning the recovery rate (5), we set *n*_rec_ equal to an average of 14 days despite strong heterogeneity in the course of disease (Guan et al., 2020). The death rate follows from (11). As we want to match the number of reported deaths from COVID-19 by *T*, the constant death rate for the period from 0 to *T* can be computed from

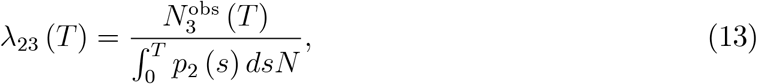

where 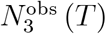 is the number of dead individuals at *T*. Employing this equation yields the value of 1*/*500 in table 2.

**Table 2.** Parameters for the epidemic without public health measures.

| $n_{\text{rec}}$ | $\lambda_{23}$ | $r$ | $\eta$ | $a$ | $\bar{\rho}$ | $\alpha$ | $\beta$ | $\gamma$ |
| --- | --- | --- | --- | --- | --- | --- | --- | --- |
| 14 | 1/500 | 0.1 | 0.4 | $3.024/10^6$ | 2/3 | 0.5751 | 0.8662 | .6459 |

A more open parameter for this epidemic but for which there is a lot of information for other epidemics is the long-run share of infected individuals, i.e. the limit 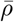 of (1). We set this equal to 0.67 meaning that once the epidemic is over, two thirds of the population will have been infected and one third are still in the original healthy state 1. As there is a lot of uncertainty concerning this value, we consider this widely employed value of 2*/*3 initially. We will then also consider one tenth of 0.67 as a lower bound further below. This lower bound is motivated by observations from Hubei and South Korea discussed in section 2.^15^ Another parameter which is hard to pin down is the share *η* of healthy individuals in state 4 (i.e. they were or are infected at some point) that can infect other individuals. We set it equal to 0.4. We also undertake robustness analyses with respect to this parameter.

The most interesting parameters are those that allow us to match data reported by RKI. To do so, we minimize the Euclidean distance between the reported data and the predicted values of the model. We target a weighted sum of the squared difference between 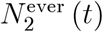 from and observation and the newly-sick 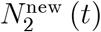 from (10) and observation. More precisely, parameters *a*, α, β and γ are obtained from

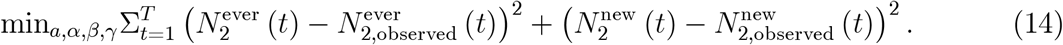

We impose constraints for α, β, γ to lie between zero and one and for *a* to be positive. None of the constraints are binding. Table 2 presents these and all the other parameter values.

We finally need to quantify *r*. A generally accepted benchmark says that around 80% of infected individuals do not display any or only weak symptoms.^16^ The remaining share of 20% turns sick. One crucial question for equation (4), but also for our long-run prediction from (12), is therefore the issue of reporting. If a large share of sick individuals show up at the general practitioner who does not test all individuals that display symptoms that might be caused by CoV-2, then the share *r* of individuals that are sick *and* reported is much lower than these 20%. We therefore set *r* = 0.1 and undertake robustness analyses further below.

#### Model for predictions

After some intermediate steps (see app. A.2), and employing the fact that we can replace probabilities in our system (7) by expected numbers of individuals as discussed in footnote 13, we obtain our final theoretical structure,

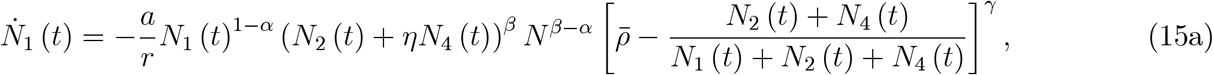

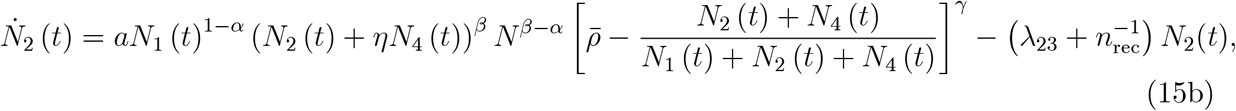

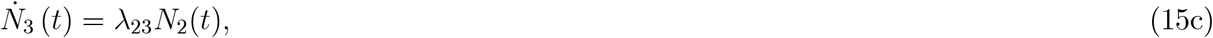

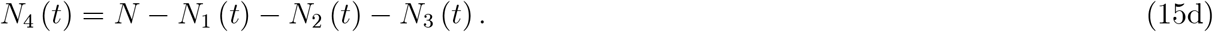

This is the system we work with to compute predictions reported in what follows. Initial conditions for our solution are *N*_3_ (0) = *N*_4_ (0) = 0 and 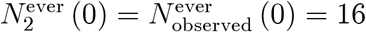 for 24 February 2020 (RKI, 2020), 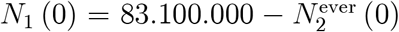 where 83.1 Million is the population size in Germany (Statistisches Bundesamt, 2020).

### 4.3 The goodness of fit of the calibration

We can judge the quality of our calibration in two ways. First, we can check the fit for observed number of individuals that were ever reported to be sick, 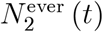 from (9). Second, we can check the quality of the match of the newly reported sick individuals, i.e. 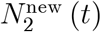 from (10).

The left part of figure 4 shows the number of individuals that were ever reported to be sick. The observations from RKI are depicted as circles. It shows that our parameter choices are sufficiently good to use this model for projections into the future.^17^ This impression is also confirmed from the right part of this figure. Obviously, fitting daily values is more demanding due to the greater variance in the data.

**Figure 4.**
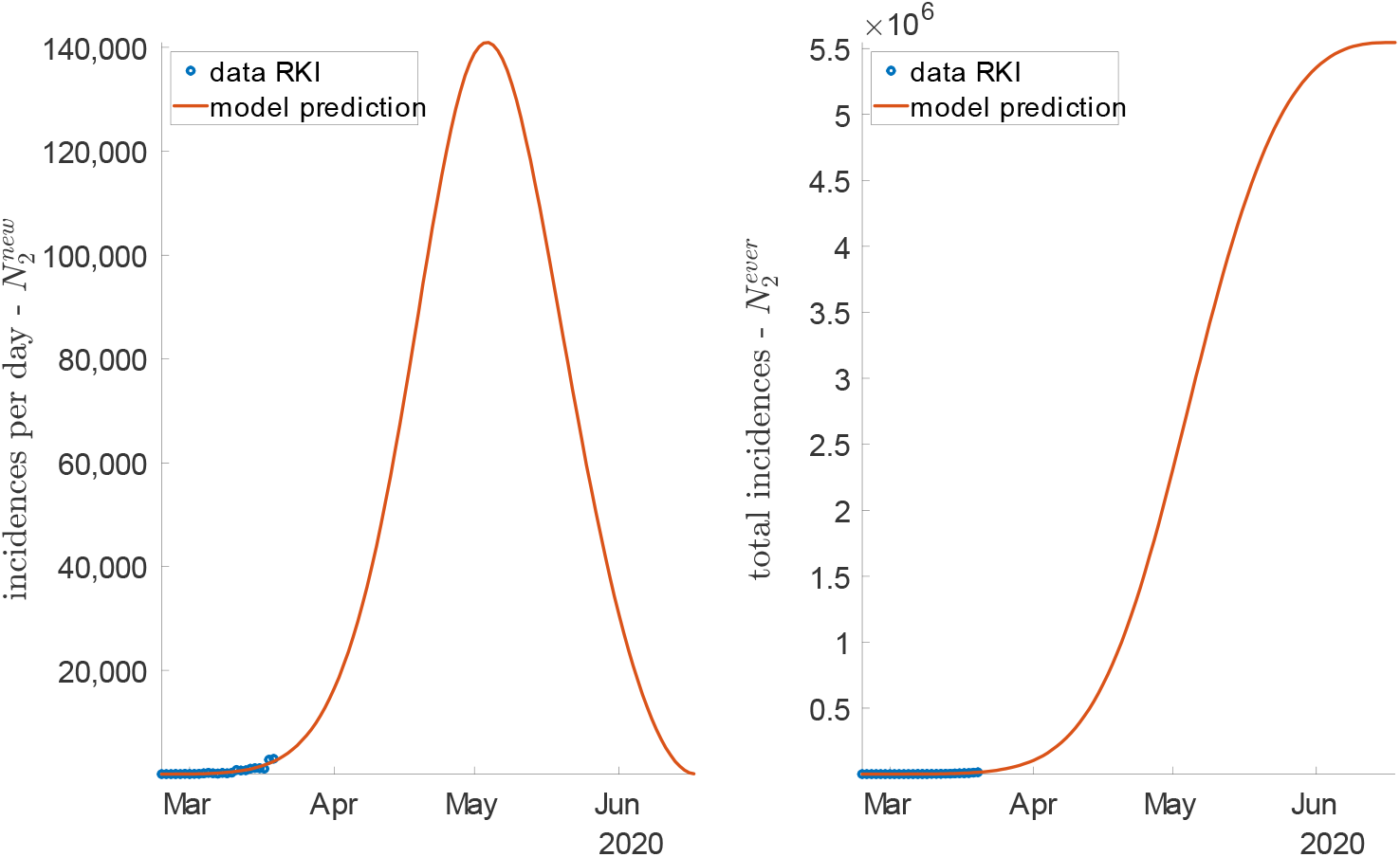
New incidences (reported) in the model (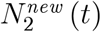, curve) and in the data (dots) (left figure) and the number 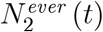 of sick in model and data (right figure) for the epidemic without public health measures

The figure also shows where, in the case of no public health measures, Germany would be heading to. Under the assumption that the share of sick *and* reported individuals out of the infected individuals is *r* = 0.1, the final number of reported sick individuals will be (right part of figure) 5.5 million. This level will be reached towards the end of June.^18^ Hence, towards the end of the epidemic, at stable parameter values, around 7 in 100 inhabitants will have been sick (and reported).

Note that this long run value does not only come out of the solution of our calibrated system (15). It can also be directly read from the long-run prediction in (12), which can be written as 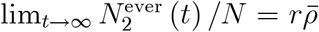 When we assume that two thirds of the population will be infected after the epidemic, 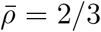, and 20% turn sick of which the half is reported such that *r* = .1, we immediately see from (12) that the long run share is 6.7%, i.e. around 7 in 100.

## 5 Findings

### 5.1 The number of sick individuals and health care

The most important quantity coming out of our analysis is probably the number *N*_2_ (*t*) of currently sick individuals. This number will decide whether hospital services will be enough for all individuals. The number of sick individuals rises strongly initially. This is the consequence of infections. As recovery sets in only after 2 weeks, the initial period sees this strong increase. Once the number of new sick cases reduces (as the number of individuals in state 1 decreases) and recovery sets in, the number of sick individuals falls again. The peak will be reached mid May. We note that this is a prediction for the case where individual contacts, as captured by the parameter *a* in (2), remain unchanged over time. If the policy measures in place as of 14 March have an effect, we would expect a slowdown in the rise of *N*_2_ (*t*). This picture therefore shows what would have happened if no interventions had taken place. In this case, the number of sick (and reported) individuals on a given day, taking recovery into account, will rise above 1 million end of April.

This figure also provides the maximum number of infected individuals over the epidemic. This allows to understand whether the number of extreme cases is always below the level that can be handled by the German health system. The peak of the number of sick individuals mid April is close to 220,000. According to the Deutsche Krankenhausgesellschaft (2020), there are 28,000 beds available in intensive care units (ICUs) in Germany. A fraction of these are currently available for the treatment of COVID-19 patients. In addition to that, an undefined number of hospital facilities allowing ventilation might be available in due time by reorganization activities. By contrast, a large fraction of COVID-19 patients might be treatable on regular wards. While it is therefore difficult to pin down the precise number of beds available for COVID-19 patients, a peak of more than 1 million on a given day is definitely beyond any level that can be handled by the health care system. The prevention of such a scenario is the key issue of all measures currently taken by federal and state governments. We therefore point to our assessment of theses measures further below.

If one would like to point out one advantage of this scenario without public health measures: the epidemic is fast. As the infection rate is very high, the outflow from the state of being healthy is fast. Hence, a large share of individuals, 1 − *r* = 90%, are infected without showing symptoms (or weak symptoms such that they are not reported) and they are immune (in state 4) quickly. Both figure 4 (left) and figure 5 show that the epidemic would be over end of June.

**Figure 5.**
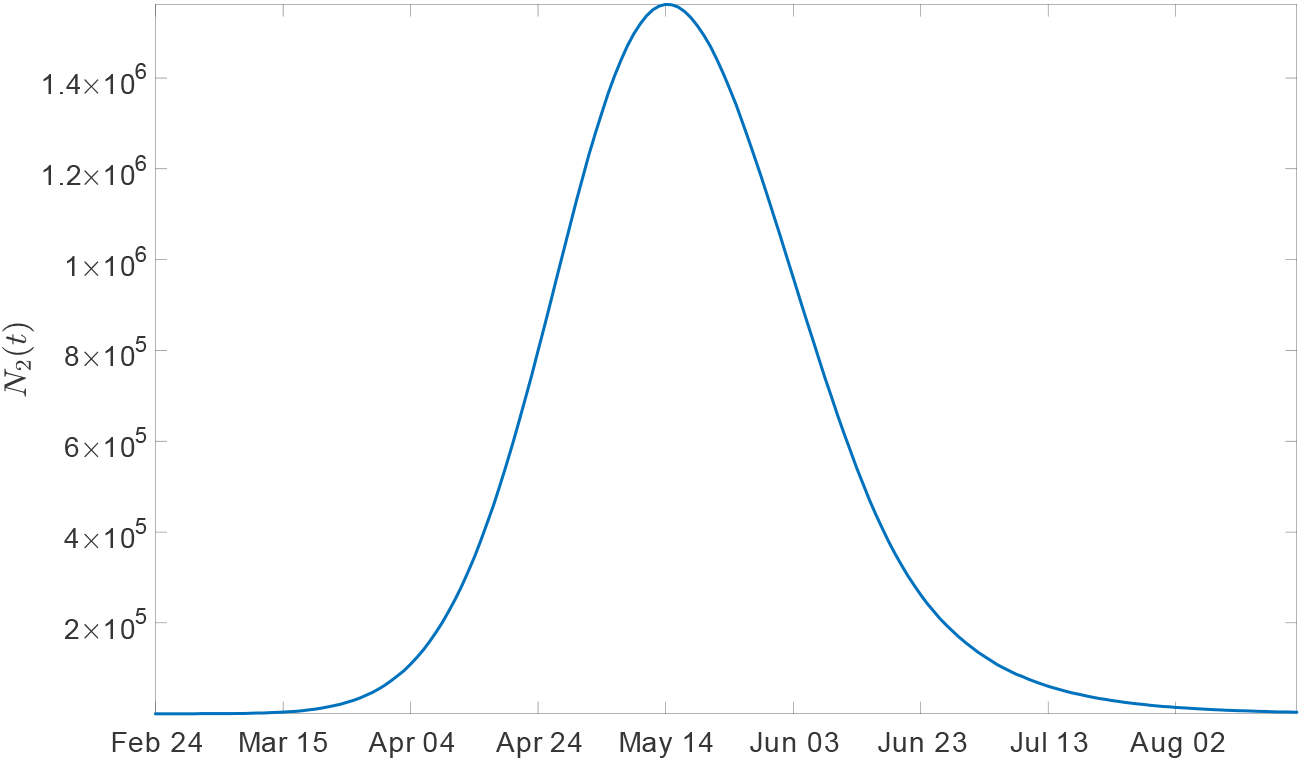
The number of sick individuals (s = 2) by day starting on 24 February 2020 in the absence of public health measures

### 5.2 Individual risks

One of the concerns of individuals is that they might get infected. How large is this risk in a world without interventions?

As this picture shows, the risk to get sick for each individual from (8) is very low throughout the entire epidemic. At the peak, the probability to get sick on a given day (red curve) is around 10^−4^. In words, 1 person in 10,000 inhabitants turns sick per day. The probability to turn sick within a week (blue curve) is of course larger and lies at the peak at around 7 in 10,000 individuals. We emphasize again that this is the number of sick individuals that are actually reported to the RKI.

We compare the daily risk of 1 in 10,000 or the weekly risk of 7 in 10,000 with the long-run risk of 7 in 100 by the end of the epidemic to be or have been sick (as discussed after figure 4), it seems that at the individual level, the daily or even weekly risk of the epidemic is not really a major health risk.^19^ It is clear, however, that at the level of society, the epidemic is a major issue. This difference between the individual risk and aggregate outcomes is the result of a classic negative externality. Even individuals that take into account the overall risk of 7 in 100 to turn sick might not be concerned. If the risk of infecting others was taken into account, individual behaviour would look differently. This is why governmental interventions have a clear justification from the perspective of market failure and public economics.

### 5.3 Robustness check

#### The long-run sickness rate *r*

Given uncertainty about the share *r* of individuals that turn sick from an infection, we undertook robustness checks. Instead of our value of 0.1, we also solved our model for an *r* ten times lower up to twice as large — as the left column of the next table shows. For each of these new values for *r*, we recalibrate the values for *a* and α to match the initial observations of sick individuals.

The model predictions are very reasonable from a qualitative perspective. The larger the individual risk *r* to get sick, the higher the number *N*_2_ (*t*) of simultaneously sick individuals and the later the peak *t*^max^ of the epidemic.

We also find, as was to be expected, that a larger *r* implies a longer duration of the epidemic. If we define the end of the epidemic by the day 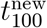where only 100 new incidences are reported per day, we see that the end of the epidemic lies between 19 May and 25 June. Interestingly, the end of the epidemic is not monotonic in *r*. When the end of the epidemic is defined by the point in time *t*_1000_ where 1000 sick individuals are left (*N* (*t*_1000_) = 1000), the epidemic ends between 15 July and 27 September. Note that the *t*_1000_ definition implies much later endpoints (of one and the same epidemic) than the 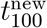definition.

Why are these numbers important from a public health perspective? The data section shows that Hubei and South Korea seem to have managed to keep the long-run ratio 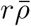 from (12) at 1 in 1000 (Hubei) or 1 in 5000 (South Korea). If we believe that these values are long-run values, then either the individual probability *r* to get sick after infection of the long-run infection rate 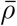 must be much lower than the standard values shown in 2 or sickness probabilities *r* must be lower.

As no information is available at this point about the share of individuals that are already immune to CoV-2 in Germany, one might well hope that Germany is heading towards a Hubei long-run equilibrium. As data quality was often argued to be an issue, our long-run ratio 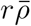 with *r* = 0.01 would correspond to 6.7 in 1000 individuals that were sick at some point in the long-run 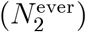. This would imply that — in the case of no public intervention — the peak in the number of sick individuals would only be around 180 thousand individuals (as the first row of table 3 shows). The blue time path of *N*_2_ (*t*) of figure 7 would be relevant.

**Table 3.** Varying the long-run sickness rate r.

| $r$ | $a$ | $\alpha$ | $\beta$ | $\gamma$ | $\max \{N_2(t)\}$ | $t^{\max}$ | $t_{1000}$ | $t_{100}^{\text{new}}$ |
| --- | --- | --- | --- | --- | --- | --- | --- | --- |
| 0.01 | $1.03 \times 10^{-4}$ | .685 | .8289 | .6952 | 182,004 | 23 April | 15 July | 19 May |
| 0.025 | $1.282 \times 10^{-4}$ | .6707 | .8397 | .6113 | 440,135 | 2 May | 4 Aug | 24 May |
| 0.05 | $12.3 \times 10^{-4}$ | .7505 | .8513 | .7921 | 775,478 | 8 May | 25 Aug | 25 June |
| 0.1 | $3.02 \times 10^{-5}$ | .5751 | .8662 | .6459 | $1,56 \times 10^6$ | 14 May | 7 Sep | 17 June |
| 0.2 | $2.21 \times 10^{-5}$ | .5481 | .8847 | .7511 | $2,9 \times 10^6$ | 19 May | 27 Sep | 16 June |

**Figure 6.**
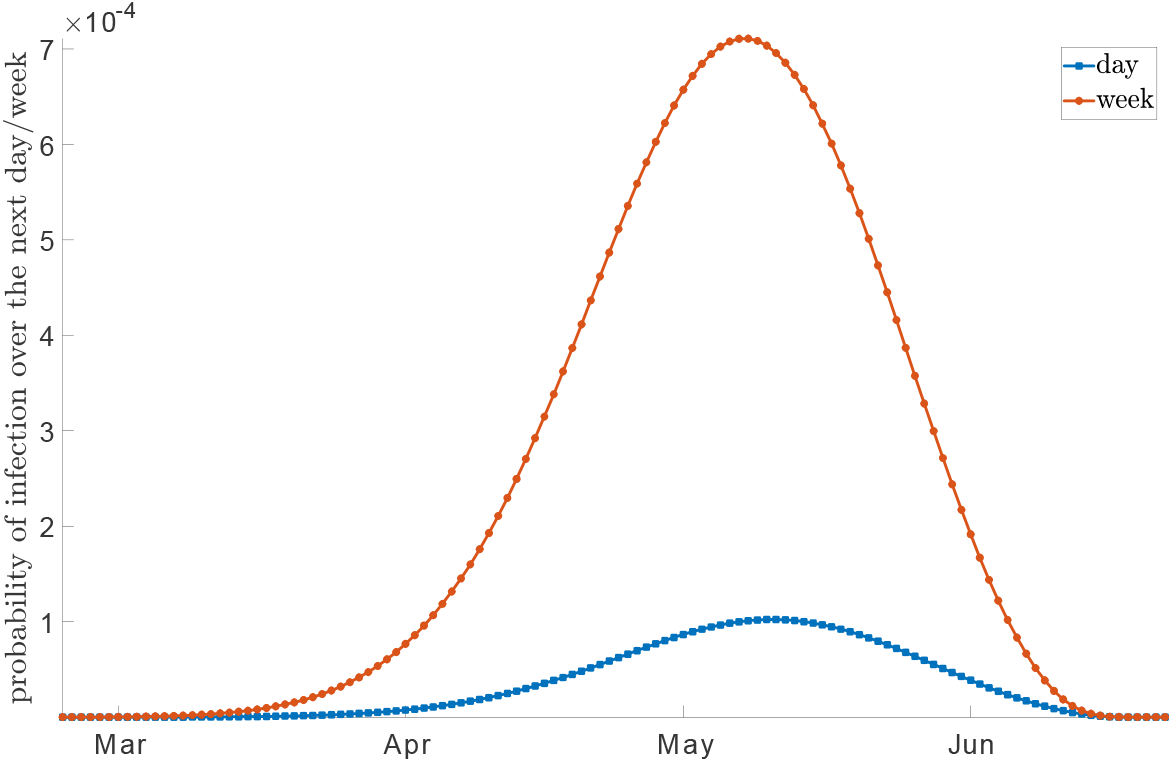
The probability per day (dotted) and per week (solid) to get sick in the absence of public health measures

**Figure 7.**
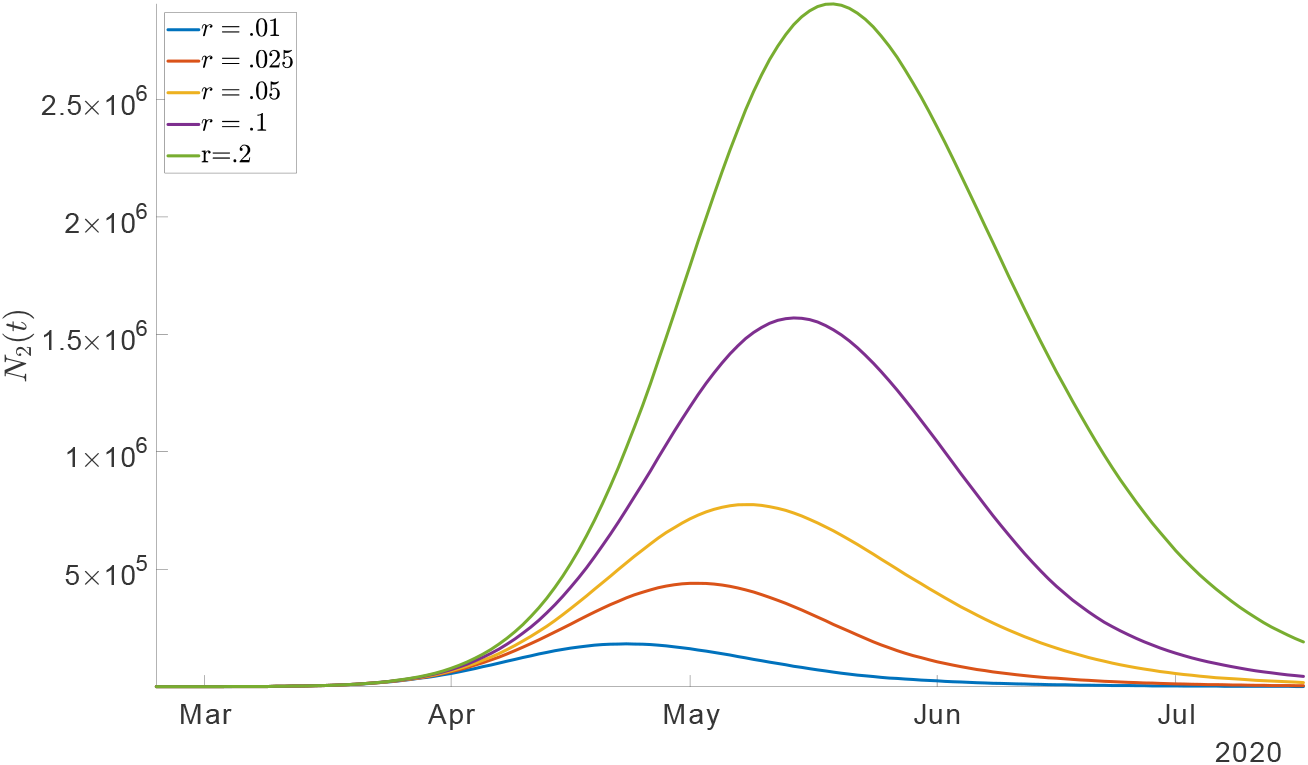
The number N_2_ (t) of sick individuals as a function of assumptions on the probability r to get sick after an infection

**Figure 8.**
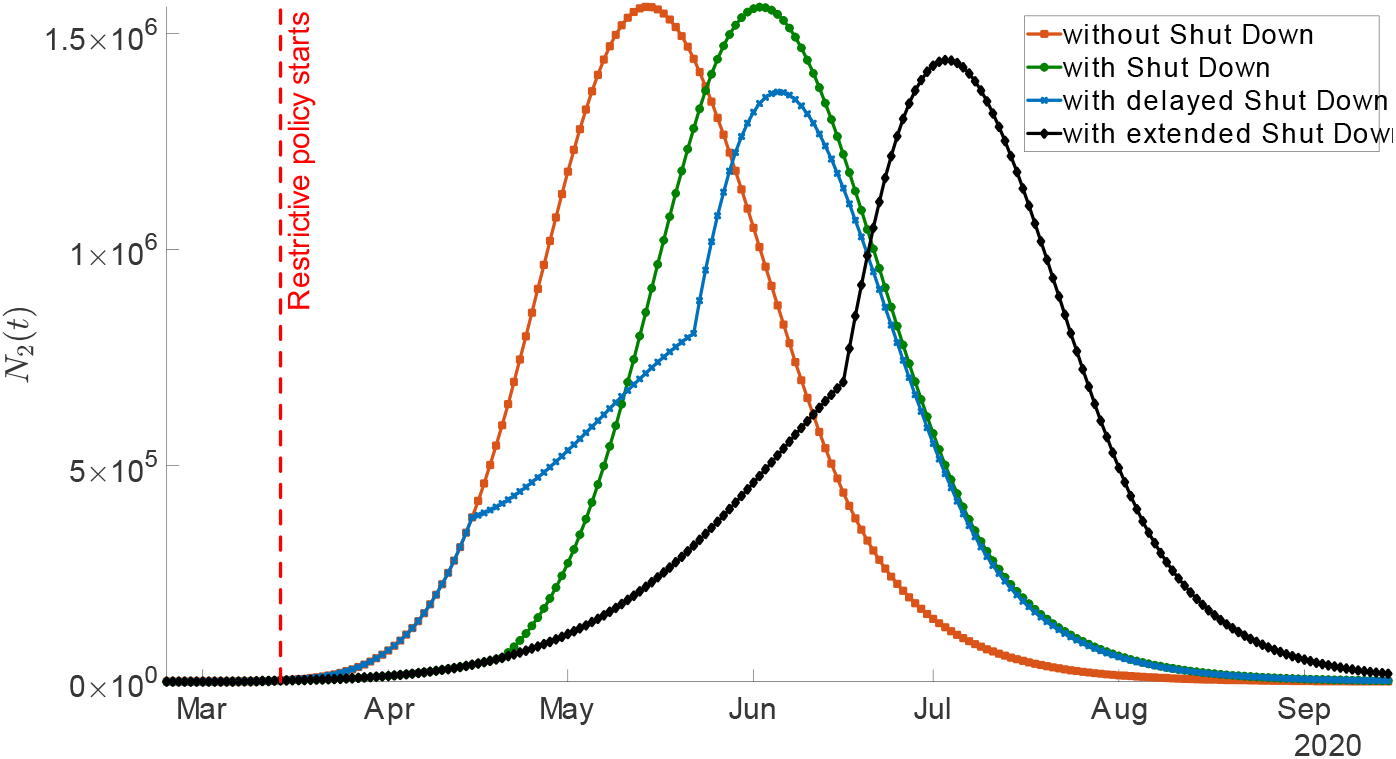
The effect of a shut down

#### Infections via individuals without symptoms

We also inquired into the effect of *η*, i.e. into the share of healthy individuals that can infect other individuals. The value of *η* in table 2 is 0.4 and we now varied it between .25 and.75. While there were qualitative changes (figures are available upon request), there are no quantitative change that would be of interest for any public health questions.

## 6 The effect of policy measures

Given that an undamped spread of CoV-2 in society leads to high reported incidences *N*_2_ (*t*), public health measures are attempting to reduce social contacts. The fewer social contacts, the lower infections per day. These contacts are captured by the parameter *a* in (2). Now assume the number of contacts per individual is restricted in some exogenous manner. Individuals are not allowed to watch soccer matches, go to conferences or move around freely in their region or country. What is the effect of these measures? We also ask whether it makes a difference whether such measures are taken at the beginning of the outbreak of infections or when the measures are delayed for 1 month.

### 6.1 The experiments

We model policy measures by assuming a time path for *a*. The first major policy measure, ‘shut down’ for simplicity, took place as of 14 March 2020. Sports events from professional to amateur leagues and schools were closed. This measure is planned to continue until 19 April 2020. As our quantitative analysis starts on 24 February, we employ parameters from table 2 for the unrestricted epidemic. We especially keep the value for the contact intensity *a* of individuals. After 20 days, we reduce *a* to a lower level *a*^low^ for the duration of this policy measure. After 37 days, i.e. on 20 April, we return to the original level, assuming that sports events and schooling resumes. Formally,

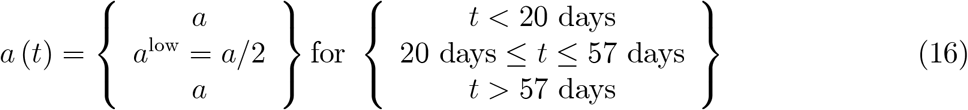

captures the shut down initiated on 13 March. For all shut down scenarios we assume that *a*^low^ is 50% lower compared to *a*.

To understand whether a later intervention is more effective, we counterfactually assume that the intervention starts 32 days later. Here we set

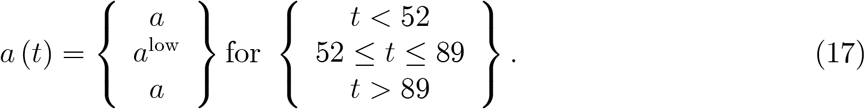

We call this the ‘delayed shut down’.

Finally, we investigate into the effect of extending the shut down. We extend the period with lower social contact rate *a*^low^ to last for 74 days, i.e.

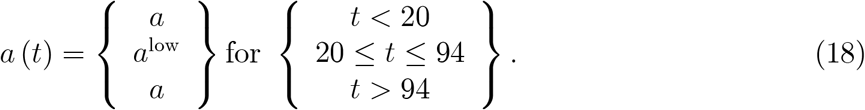

This is our ‘extended shut down’.

### 6.2 The quantitative effects of policy measures

#### Baseline analysis

For this baseline analysis, we do not recalibrate the model as *a* in our scenarios changes after *T*, the day providing the last observation used for calibration. Parameter values are therefore given by the values in table 2. The effect of various forms of shut downs are captured by a drop of the contact rate *a* by 50% to *a*^low^, as shown in (16).

The results are shown in the next figure. Our main variable for judging policy measures is the number *N*_2_ (*t*) of individuals that are simultaneously sick.

First, the policy is effective. Reducing contacts reduces the speed of infections and new cases of sickness — but also the transition from healthy in state 1 to healthy (with infection) in state 4. The case without shut down is illustrated by the red curve in this figure. It is identical to the curve in figure 5. The shut down from (16) which reflects the planned closing of schools in Germany until mid April is drawn as the green curve. It reduces the rise of the number of sick individuals until mid April. Afterwards, the number of sick individuals will rise more quickly again. The peak is reached in June and this peak is lower than the one without the shut down. In this respect the shut down is a good policy measure. When the shut down is extended (the black curve), the peak is further shifted to the right and also smaller.^20^

Quantitatively speaking, the shut down prolongs the epidemic in terms of its end as defined by 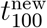 also employed in table 3 (100 or fewer new incidences are reported per day). For the shut down currently in place and for the delayed shut down, the end is beginning of July. For the longer shut down, the end is beginning of August. Concerning the peak demands on the health system, the shut down as currently in place, and also an extended shut down, hardly have any effect. There is a certain delay — which helps the health sector to prepare for higher numbers in the future. But the level of the peak, as the figure shows, hardly changes. This looks like a bleak outlook.

To be fair, we should stress at this point that the assumption that a shut down reduces the contact rate and thereby the infection rate by 50% is open to empirical investigation. At this point, this is a model assumption for which empirical evidence is still lacking. Hartl et al. (2020) show that statistically convincing evidence will be available at the earliest beginning of April. It seems fair to say, however, that it can hardly be expected at this point that current measures reduce the infection rate by 50%. Hence, the above evaluation of the shut down might even be too optimistic.^21^

#### Is a delayed shut down a good idea?

Interestingly, a delayed shut down (blue curve) is almost as good as an extended shut down and in some respect is even better: A delayed shut down reduces the peak in the number of sick individuals. Hospitals are therefore less crowded with a delayed shut down as opposed to an immediate shutdown. This implies, more generally speaking, that there is an optimal timing for a shut down.

Why is there an optimal point for a shut down? Imagine the shut down takes place early (our curve). Then the number of sick individuals does not increase that quickly. The downside is that, once the shut down is over, there are a lot of healthy individuals left that can turn sick. The number of new incidences jumps up and the peak is reached quickly. When the shut down takes place later (the blue curve), the number of sick individuals is larger at the moment the shut down starts. Once the shut down is over, the number of healthy individuals that are left to turn sick is not as large as in the case of an early shut down. As a consequence, the peak is less high.

In a perfect world with complete knowledge about the number of sick individuals, the long run infection rate 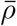, the individual risk *r* to turn sick from an infection and other parameters employed in this model, policy makers should therefore choose the optimal point in time for an intervention. In the world we live in, this information is not available, neither from a medical, nor from a statistical and data processing perspective. The finding nevertheless makes the strong point that policy interventions should be conditioned on the current situation in a country or region. Ideas that policy measures must be the same for all regions in Germany at all points in time are strongly contradicted.^22^

#### The policy effects with lower 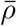

Let us now return to the uncertainty about the value of the long-run share 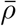 of infected individuals. We varied *r* in table 3 such that we could also study a “data-quality adjusted Hubei-scenario” where 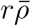 implies that there are 6.7 in 1000 (ever) sick individuals. This was discussed after figure 7. We now study changes in 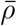 at unchanged *r* = 0.1 such that we end up at 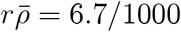 We therefore reduce 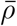 by a factor of 10. We do recalibrate our model for this scenario as 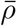 affects the calibration outcome. The fit is similarly good to the one discussed after figure 4. Given the new parameter values, we change the contact rate according to the shut down (16) and the extended shut down (18).

Let us start with the long-run effects. As we know from (12), the long-run effects with 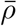 reduced by a factor of 10 are 10 times lower. This is visible in the right part of figure 9 displaying 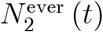 Once the epidemic is over, there will be 550 thousand individuals that were sick at some point, one tenth of the level in figure 4. This is again independent of policy measures as the latter do not affect 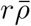.

**Figure 9.**
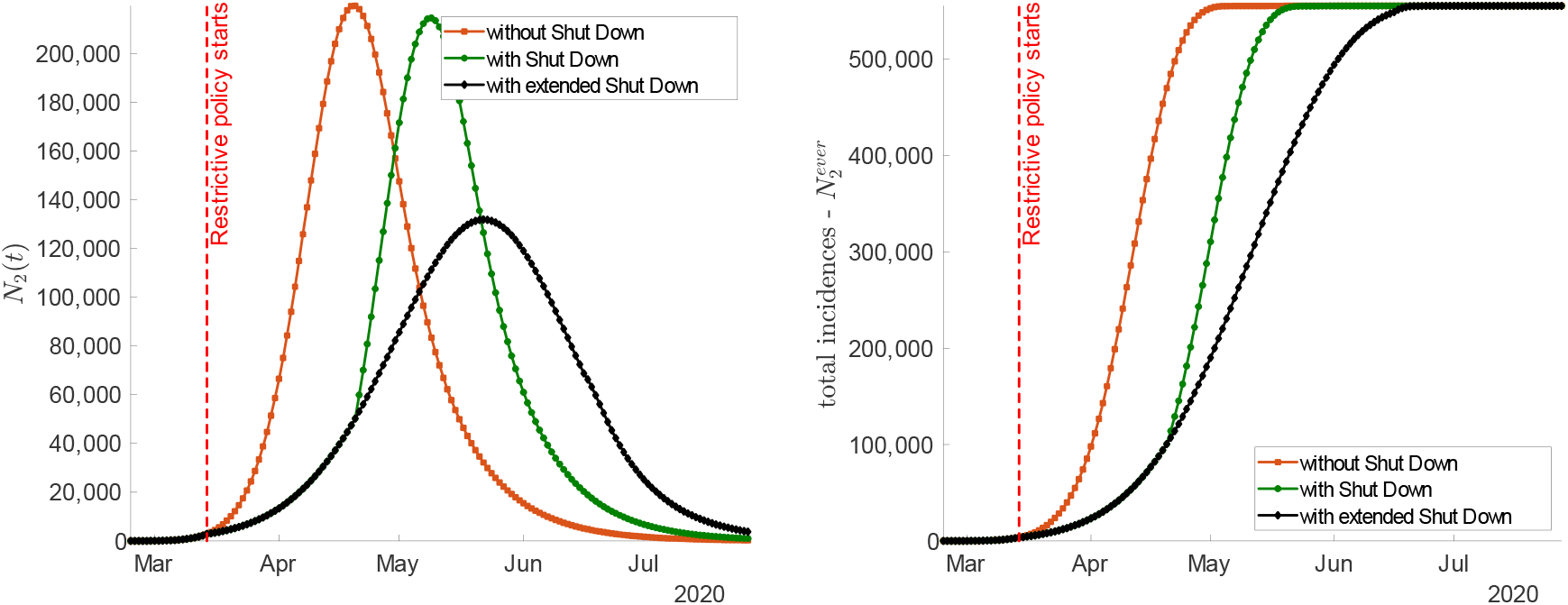
The effect of changes in the long-run infection rate 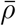

**Figure 10.**
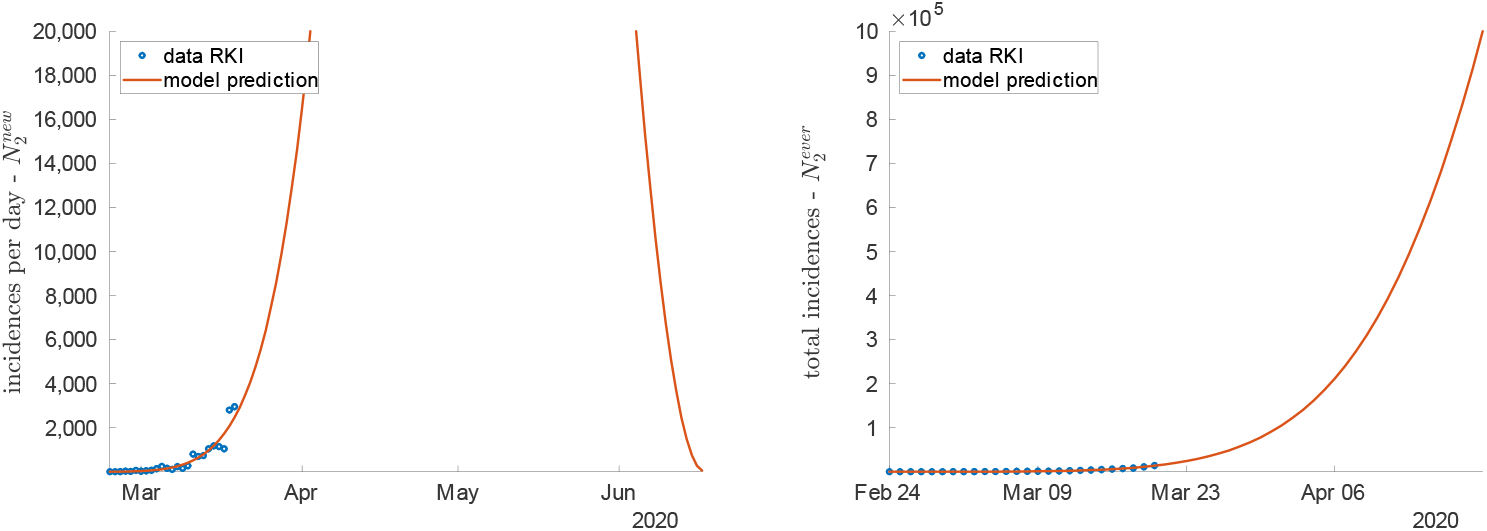
Enlarged version of figure 4 with data and model fit

Concerning short-run effect, we see that in the case of no intervention (or interventions that do not have an effect), the peak comes earlier for *N*_2_ (*t*). The red curve in figure 9 peaks in mid April in contrast to early to mid May for the higher long-run infection rate of figure 8. The effect of the shut down currently in place looks qualitatively similar to the effect when 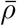 is higher. By contrast, the extended shut down in figure 9 does not display a strong rise in growth rates of *N*_2_ (*t*) once it is over. It is directly followed by the decline in the number of sick individuals.

Most importantly, however, theses two figures display a strong quantitative difference. If the long run infection rate in Germany is in the “data-quality adjusted Hubei range”, the peaks are at a much lower level. In the case without policy measures and in the case where the current shut down (16) is the only policy measure, the peaks of the simultaneously sick number of individuals is just above 200 thousand. This strongly contrasts to earlier values above 1 million. Another difference consists in the quantitative difference here between the extended shut down and the shut down. In the scenario with a lower long-run infection rate, an extended shut down implies that the peak of *N*_2_ (*t*) will be much lower than the peak of the normal shut down. This was not the case in figure 8. This points to the fact that shut downs need to be sufficiently strong relative to the overall potential number of sick individuals.

When the long-run infection rate is in this Hubei-range, then the epidemic is also over more quickly than in the case of an infection rate of two thirds. In any case, the epidemic will last until June, in the case of extended shut downs (or other measures) it can even last until mid July.

## 7 Conclusion

We have developed a model that allows to study the spread of an infection in a society. We have solved this model and calibrated it to Germany. We employ the observed number of reported infections to match the initial increase of the number of sick individuals. In addition, we employ parameter values from the medical literature to quantify e.g. long-run infection rates or individual risks to turn sick after an infection. Given uncertainty about the precision of these parameter values, we undertook robustness checks with respect to this long-run measure.

When we studied the effect of policy measures, we found that any intervention that reduces the contact rate of individuals reduces the strength of the epidemic. Both the peak of the number of simultaneously sick individuals as well as the individual sickness risk go down. This decrease is stronger, the longer the policy measure lasts.

We also find, however, that there is an optimal point in time when an intervention, effective for a fixed length of time (say, 3 weeks), should start. When the intervention starts too early, there are “too many” healthy individuals left at the end of the intervention. The rise in the number of sick individuals would be accelerated. When the intervention starts later, the number of sick individuals is larger initially. Once the intervention is over, there are, as a consequence, not so many healthy individuals left and the peak is smaller. While implementing optimal schemes for real world societies probably requires too much information, this finding points towards implementing policy measures which are contingent on the current situation (number of sick individuals per capita) of a region or country.

In the long-run, for given parameters, the number of sick individuals is not affected by policy measures when the latter only reduce contact rates of individuals. All findings are subject to large uncertainty given little knowledge about long-run infection rates following a CoV-2 epidemic and individual sickness rates. It seems to be certain, however, that the epidemic will last at least until July, given the public health measures in place. Even in the case of a good scenario with extremely low long-run infection rates, the peak of the number of simultaneously sick individuals will lie above 200 thousand individuals.

A lot of extensions of this framework are worth being undertaken. First, optimal behaviour of individuals could be taken into account. The activity parameter *a* is like the outcome of decisions of individuals on how much to work and how much to enjoy social interactions. One can imagine a trade-off between reducing the risk of getting infected by lowering *a* and working fewer hours and thereby experiencing lower labour income. The same trade-off is present at the aggregate level to be tackled by a government. Such a trade-off might also include behavioural features like anxiety as in Caplin and Leahy (2001), potentially in a simplified way as presented in Wälde and Moors (2017). These extensions will help to better understand the challenges posed by this pandemic.

A more detailed modelling of the long-run infection rate would also be useful. The path of social contacts over time should have an influence on the share of infected individuals in the long-run. A more detailed matching mechanism than the one employed here — borrowed in spirit from search and matching models — should be developed.

Finally, the calibration process employed here can be extended to full structural estimation. Further, the effects of the policy measures in place since 14 March can be estimated or calibrated. The calibration in this paper employs data up to the point where measures of 14 March should start having an effect. As of end of March, the intervention regime discussed above and the corresponding (reduced) contact rate could be quantified. This would provide a good idea of the effects of these policy measures.

## Data Availability

All of the data used has been collected from the WHO situation reports, the JHU COVID19 dashboard maintaining by Dong et al (2020), and Eurostat for the population statistics.

https://www.who.int/emergencies/diseases/novel-coronavirus-2019/situation-reports

https://www.arcgis.com/apps/opsdashboard/index.html#/bda7594740fd40299423467b48e9ecf6

https://ec.europa.eu/eurostat/tgm/refreshTableAction.do?tab=table&plugin=1&pcode=tps00001&language=en

## A Appendix

### A.1 Data sources

#### Data sources and calculations

The data used in this note is taken from two sources: (i) the World Health Organisation’s Situation reports on the progress of COVID-19, and (ii) the dataset put together by the nCoV-2019 Data Working Group (Dong et al. (2020)). In particular, wherever available the data from the nCoV-2019 Group is used instead of the data from the WHO, in an effort to remain as consistent as possible by using only one source at a time. Data from the nCoV-2019 Group is used as it provides a centralised and standardised repository of multiple official sources.

The figures showing the number of new confirmed cases at a date *t* were produced by computing the difference between the cumulative total number of confirmed cases at *t* minus the cumulative total number of confirmed cases at *t* − 1, i.e.

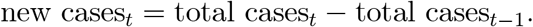

As detailed in the Table 1, the infection rate is computed by dividing the cumulative total number of confirmed cases as of the time of writing by the population of the corresponding country in 2019. Meanwhile, the risk factor is simply the inverse of that fraction and is given by the total population divided by the cumulative total number of confirmed cases, i.e.

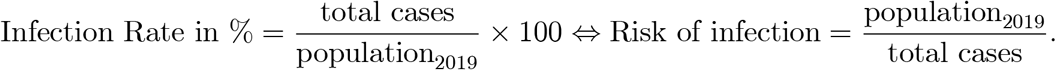

### A.2 Intermediate steps for (15)

Recall (2) and write it as

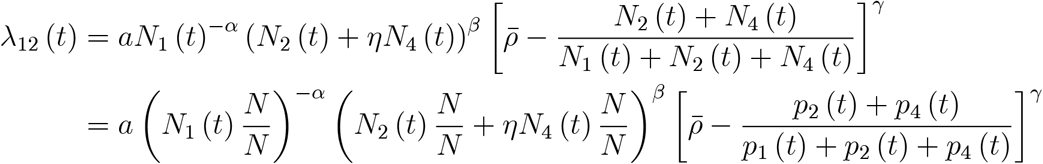

where the fraction in squared brackets was multiplied by *N/N* as well and we employed *p*_*s*_ (*t*) = *N*_*s*_ (*t*) */N*. Applying this to term one and two as well,

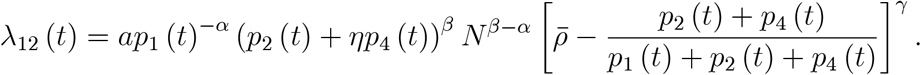

If we want homogeneity of degree 0, we would set α = β. Employing the transition rate λ_14_ from (4) and taking λ_24_ from (5) into account, our model in (7) becomes

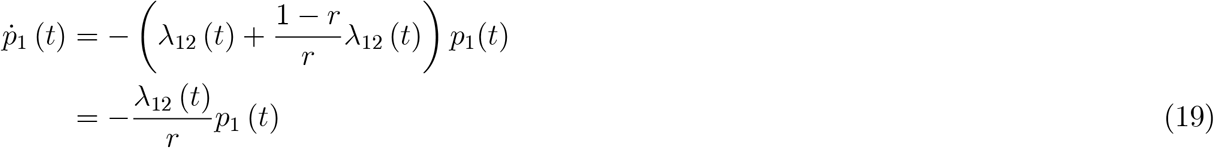

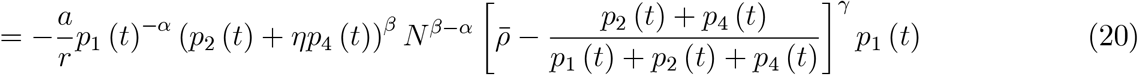

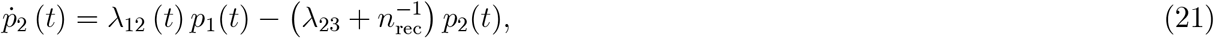

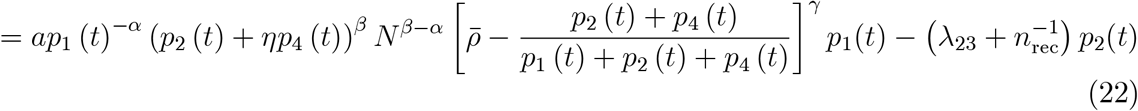

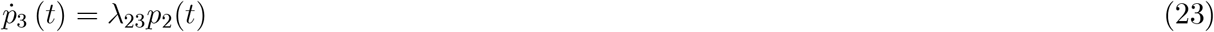

Simplifying, we get (15) in the main text.

### A.3 Goodness of fit

We provide a graphical illustration of how well our parameter choice via the minimization procedure in (14) fits the data.

## Footnotes

2 An essay by Simon (2020) argues that the death rate in Germany is much smaller than in other countries. This makes it difficult to apply the Ferguson et al. (2020) findings to Germany.

3 There are also various internet pages that offer projections for parameter models that can be chosen by the user. Examples include https://gabgoh.github.io/COVID/index.html, https://neherlab.org/covid19/ or http://covidsim.eu/. These pages usually do not show the details of the model and a comparison to our approach is therefore difficult.

4 Eichenbaum et al. (2020) study the effects of the CoV-2 epidemic on economic decisions of households. They emphasize the household decisions to reduce labour supply exacerbate the recessionary effects of the epidemic. They apply their model to the US. Barro et al. (2020) employ mortalities and economic consequences of the “Spanish Flu” to provide estimates of the world wide consequences of COVID19. They do not offer a detailed projection for Germany. The costs for Germany are estimated by Dorn et al. (2020). Atkeson (2020) provides esitmates of economic consequences of COVID19 for the US.

5 When working with continuous states like e.g. wealth or labour income, partial differential equations result. See e.g. Bayer and Wälde (2010a), Achdou et al. (2020), Kaplan et al. (2018) or Khieu and Wälde (2020).

6 Note that all figures below are as of 26 March 2020 and come from the Johns Hopkins University dataset from Dong et al. (2020).

7 There are attempts to correct supposedly under reported numbers from China by employing South Korea as a benchmark country (Lachmann, 2020).

8 In our quantitative solution below, the initial state follow from the number of reported sick individuals on 24 February 2020.

9 Individuals that have died no longer play a role in social interactions. Hence, *N*3 (*t*) does not appear as argument in the sickness rate.

10 A parallel can be drawn between φ and a matching function, and λ12 and the job finding probability in search and matching frameworks in the tradition of Diamond (1982), Mortensen (1982) and Pissarides (1985).

11 We employ “around” as some individuals will have ended up in state 3 whose number does not enter the expression in (1).

12 One could plausibly argue and find support in micro studies that the death and recovery rate also depend on the number of healthy individuals (that are required to take care of sick individuals). These considerations are not crucial for our macro analysis and are therefore neglected.

13 Let us denote the expected number of individuals in state *s* at *t* by 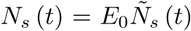 where 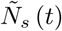 is the stochastic process for the number of individuals in state *s* at *t*. As the expected share of individuals in state *s* equals the probability to be in this state, *N*_s_ (*t*) */N* = *p*_s_ (*t*), we can write *N*_s_ (*t*) = *p*_s_ (*t*) *N* for the expected number of individuals in this state. Replacing the probabilities *p*_s_ (*t*) by *N*_s_ (*t*) would yield an ODE system in *N*_s_ (*t*). Our system employed for predictions below in (15) is expressed in this way.

14 See https://www.macro.economics.uni-mainz.de/corona-blog/ for more details.

15 One might conjecture to employ so-called reproduction numbers to get an idea about 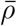 A reproduction number gives the number of infections in a non-immune population when one infected individual is introduced. Riou and Althaus (2020) estimate this value to lie in the range of 2.0 to 2.5 for Wuhan from December 2019 to January 2020. The WHO (2020) reports it in the same range. These reproduction numbers change over time (as our sickness rate λ12 (*t*) changes over time) when the population becomes more and more immune. One can therefore not draw conclusions from these estimates about long-run values like 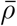. We return to this point when discussing the robustness of findings with respect to *r* after figure 7 and the effects of policy measures in figure 9.

16 This percentage is from findings in Wuhan/ China, see Wu and McGoogan (2020).

17 Appendix A.3 provides an enlarged version of this figure.

18 This figure intuitively shows that in the medium-run, the long-run sickness share 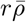 from (12) will be predictable from these observed time paths. At this point, there are too few observations and this is not yet feasible.

19 This might explain behaviour of some individuals who seem to ignore governmental pleas to reduce social interactions.

20 If we had a sequence of shut downs with breaks in between, it would qualitatively look like a combination of the green and the blue curve. The green curve represents a shut down that ends end of April where the green curve starts rising quickly again. Then when a new shut down takes place as of mid May, the number of sick individuals would follow the blue curve. When the “blue shut down” ends end of May, the number of sick individuals starts rising again as shown by the blue curve.

21 All of our scenarios could look much more optimistic if we had a vaccine. To the best of our knowledge, this will not be available over the next 6 months.

22 We understand that from a policy perspective it is simpler to “sell a measure” that is the same for everybody than measures that differ across individuals. Yet, treating unequals as equals has rarely been a convincing idea.

## Notes

### Competing Interest Statement

The authors have declared no competing interest.

### Funding Statement

No external funding was provided.

